# Analyzing the U.S. Post-marketing safety surveillance of COVID-19 vaccines

**DOI:** 10.1101/2021.07.10.21260304

**Authors:** Omar M. Albalawi, Maha I. Alomran, Ghada M. Alsagri, Turki A. Althunian, Thamir M. Alshammari

## Abstract

**Background:** Since December 2020, three COVID-19 vaccines have been authorized in the United States (U.S.) and were proceeded by large immunization programs. The aim of this study was to characterize the U.S. post-marketing safety (PMS) profiles of these vaccines with an in-depth analysis of mortality data.

**Methods:** This was a retrospective database analysis study. Details of the U.S. PMS reports (15 December 2020 to 19 March 2021) of the three vaccines (Pfizer-BioNTech, Moderna, and Janssen Ad26.COV2.S) were retrieved from the U.S. Vaccine Adverse Event Reporting System (VAERS). A descriptive analysis was conducted to characterize the reported adverse events (AEs). A comparative (Pfizer-BioNTech vs. Moderna) analysis of mortality was conducted. The mean count ratio of death between the two vaccines was estimated using a negative binomial regression model adjusting for the measured confounders.

**Results:** A total of 44,451 AE reports were retrieved (corresponding to 0.05% of the U.S. population who received at least one dose). The most commonly reported AEs were injection site reactions (30.4% of the reports), pain (reported in 26.7% of the reports), and headache (18.6% of the reports). Serious AEs were reported in only 14.6% of the reports with 4,108 hospitalizations. The total number of deaths was 1,919 with a mean count ratio of Moderna (n=997) vs. Pfizer-BioNTech (n=899) of 1.07 (95% confidence interval 0.86 to 1.33).

**Conclusions:** The vast majority of PMS AEs in the U.S. were non-serious, and the number of serious AEs is very low given the total number of vaccinated U.S. population.

## 1. Introduction

The outbreak of severe acute respiratory syndrome coronavirus disease 2 (known as COVID-19) was first reported on 31 December 2019, in China and has become a global pandemic [1]. As of 19 March 2021, more than 29.71 million cases were identified in the United States (U.S.) with a total number of death cases of at least 540,733 [2]. The first COVID-19 vaccine (Pfizer-BioNTech) authorized for the use in the U.S. on 11 December 2020 [3], and was followed by the authorization of two other vaccines: Moderna [4], and Janssen Ad26.COV2.S [5]). All these three vaccines authorized under the Emergency Use Authorization (EUA) framework (i.e. accompanied with a degree of pre-marketing safety uncertainty) [6]; thus necessitating an extensive post-marketing safety (PMS) assessment in the context of pharmacovigilance activities.

In Phase III clinical trials, the most commonly identified non-serious AEs related to these vaccines were pain at the injection site, fatigue, headache, myalgia, chills, arthralgia, and fever [7–9]. These findings were also commonly reported after the U.S. Food and Drug Administration (FDA) authorization of Pfizer-BioNTech and Moderna vaccines in addition to dizziness and Nausea [10]. In Phase III clinical trials, the incidence of serious AEs was low and comparable between the vaccines and placebo groups (0.4 to 0.6%) [7–9]. The number of death cases were lower in the three vaccine groups compared to placebo groups (2 to 6 vs. 4 to 16; respectively) [7–9]. In the first US PMS report, serious AEs were reported in 640 reports, including 113 death cases, after COVID-19 vaccinations [10]. Given the degree of uncertainty associated with the safety profile of these vaccines due to their approved under EUA, we aimed at evaluating the PMS of the current COVID-19 vaccines authorized for use in the U.S. in the context of spontaneous reporting with an in-depth analysis of the mortality data.

## 2. Methods

### 2.1 Data source

This retrospective analysis was conducted using the publically available database of the U.S. Vaccine Adverse Event Reporting System (VAERS). VAERS is a critical component of the national passive surveillance (spontaneous reporting) system of the approved vaccines in the U.S. [11]. VAERS was established in 1990 under the joint administration of the Centers for Disease Control and Prevention (CDC) and the U.S. FDA [12]. The purposes of VAERS have been to detect early warning safety signals and to generate hypotheses about possible new unexpected AEs or changes in the frequency of the known ones [11,12]. Reports in VAERS are sent from manufacturers, healthcare professionals and the public [11]. It has been a source of data for many published studies[12–15].

Data were extracted from 15 December 2020, to 19 March 2021. Data (available in tables) were extracted, assessed and cleaned by of the research team. Demographics (e.g., person, place, and time), onset intervals, and potential confounders in the mortality assessments (cardiovascular diseases [CVD], respiratory diseases, diabetes, and renal diseases) were identified in addition to the history of allergies or allergic. AEs were classified as serious AEs defined by the federal regulatory[16].

### 2.2 Statistical analysis

Distributions of non-serious and serious AEs per state, gender, vaccine, age group, and time of AE were summarized. These findings were summarized using a population-level estimate (i.e. based on the number of people who received at least one dose of the study vaccine in the U.S up to 19 March 2021 as a denominator), and using the reporting rate (i.e. using the number of reports per 100,000 doses as a denominator). From 14 December 2020 to 19 March 2021, 85,778,745 persons received at least 1 dose of COVID-19 with a total of 118,313,818 doses in the U.S. [17]. Of these, 59.21 million doses were for Pfizer-BioNTech COVID-19 vaccine, 56.94 million doses were for Moderna vaccine, and 2.03 million doses for Janssen Ad26.COV2.S vaccine [18].

An in-depth, comparative analysis of mortality between Pfizer-BioNTech vs. Moderna was conducted with the mean count ratio as a primary effect estimate (Janssen Ad26.COV2.S was excluded from the analysis given its recent approval). The mean count ratio of death between the two vaccines was estimated using a negative binomial regression model adjusting for the measured confounders (age, gender, history of allergy, onset intervals after the first dose, and the presence of several chronic diseases [Cardiovascular disease, diabetes, a respiratory disease, and renal disease]). The negative binomial model was used instead of a poisson model due to the presence of overdispersion. All analyses were completed using RStudio Version 1.4.1103.

## 3. Results

By 19 March 2021, 44,451 AE reports (0.05% of the total 85,778,745 vaccinated U.S. population) were extracted from VAERS. The median age in the identified reports was 49 years (range 13-115 years), and most symptoms occurred within two days of the vaccination (range 0-64 days). More than two-third of patients were female, and less than one-third have a type of allergy (Table 1). The most frequently reported AEs (per total number of AE reports) were injection site reactions (13,480 [30.3%]), pain (11,897 [26.7%];), headache (8,271 [18.6%]), chills (6,347 [14.0%]), fatigue (5,972 [13.0%]), and pyrexia and dizziness (4,611 [10.3%] and 4,537 [10.0%], respectively) (Table 1). Of the 44,451 AE reports, 23,890 (53.7%) were reported after the vaccination with Pfizer-BioNTech., 19,424(43.7%) after the vaccination with Moderna, and 1,107 (2.5%) after the vaccination with Janssen Ad26.COV2.S (Table 2).

**Table 1.**
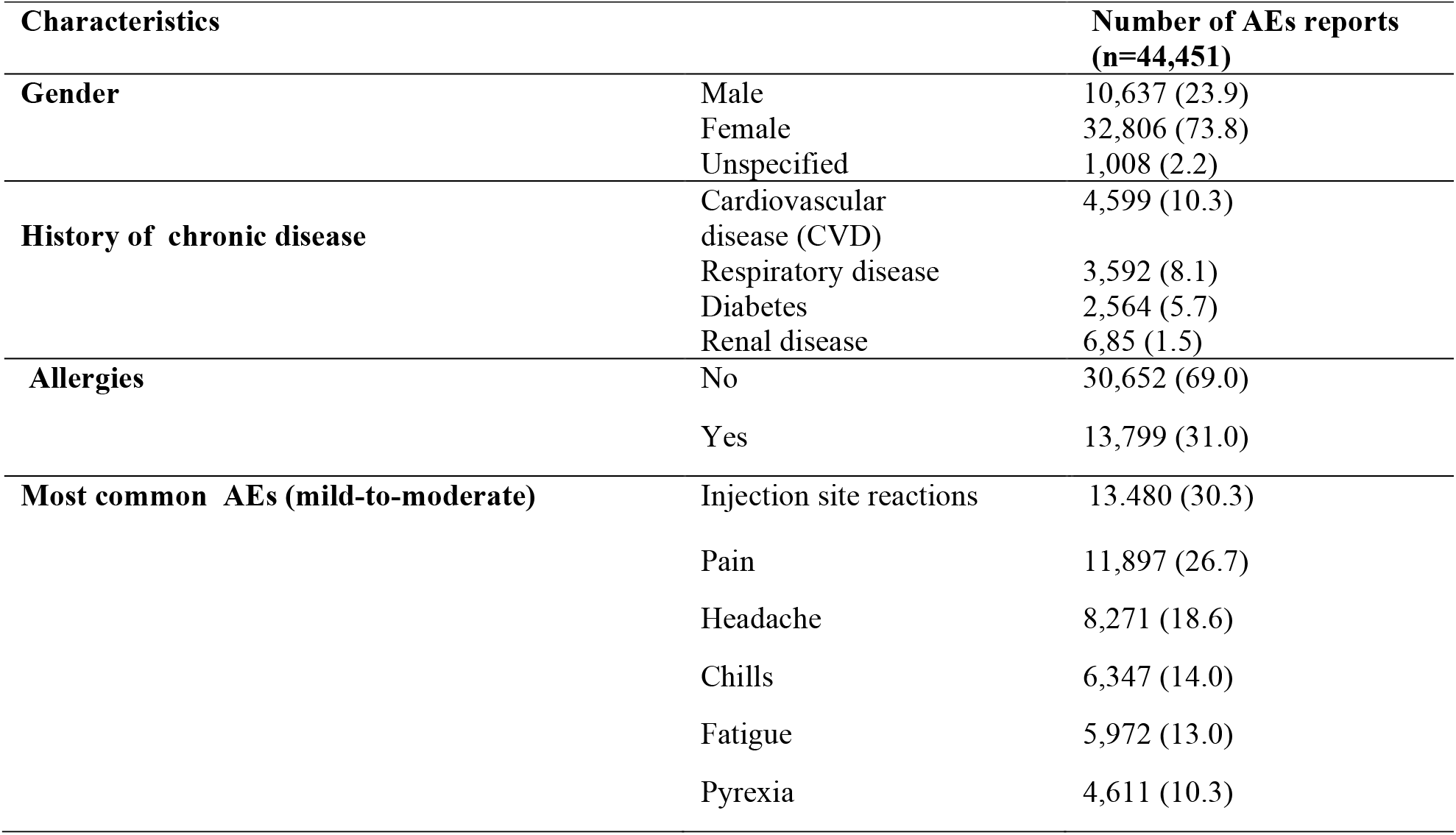
Characteristics of reported cases of AEs after receipt of COVID-19 vaccines

| Characteristics |  | Number of AEs reports<br>(n=44,451) |
| --- | --- | --- |
| **Gender** | Male | 10,637 (23.9) |
|  | Female | 32,806 (73.8) |
|  | Unspecified | 1,008 (2.2) |
| **History of chronic disease** | Cardiovascular disease (CVD) | 4,599 (10.3) |
|  | Respiratory disease | 3,592 (8.1) |
|  | Diabetes | 2,564 (5.7) |
|  | Renal disease | 6,85 (1.5) |
| **Allergies** | No | 30,652 (69.0) |
|  | Yes | 13,799 (31.0) |
| **Most common AEs (mild-to-moderate)** | Injection site reactions | 13,480 (30.3) |
|  | Pain | 11,897 (26.7) |
|  | Headache | 8,271 (18.6) |
|  | Chills | 6,347 (14.0) |
|  | Fatigue | 5,972 (13.0) |
|  | Pyrexia | 4,611 (10.3) |

**Table 2.**
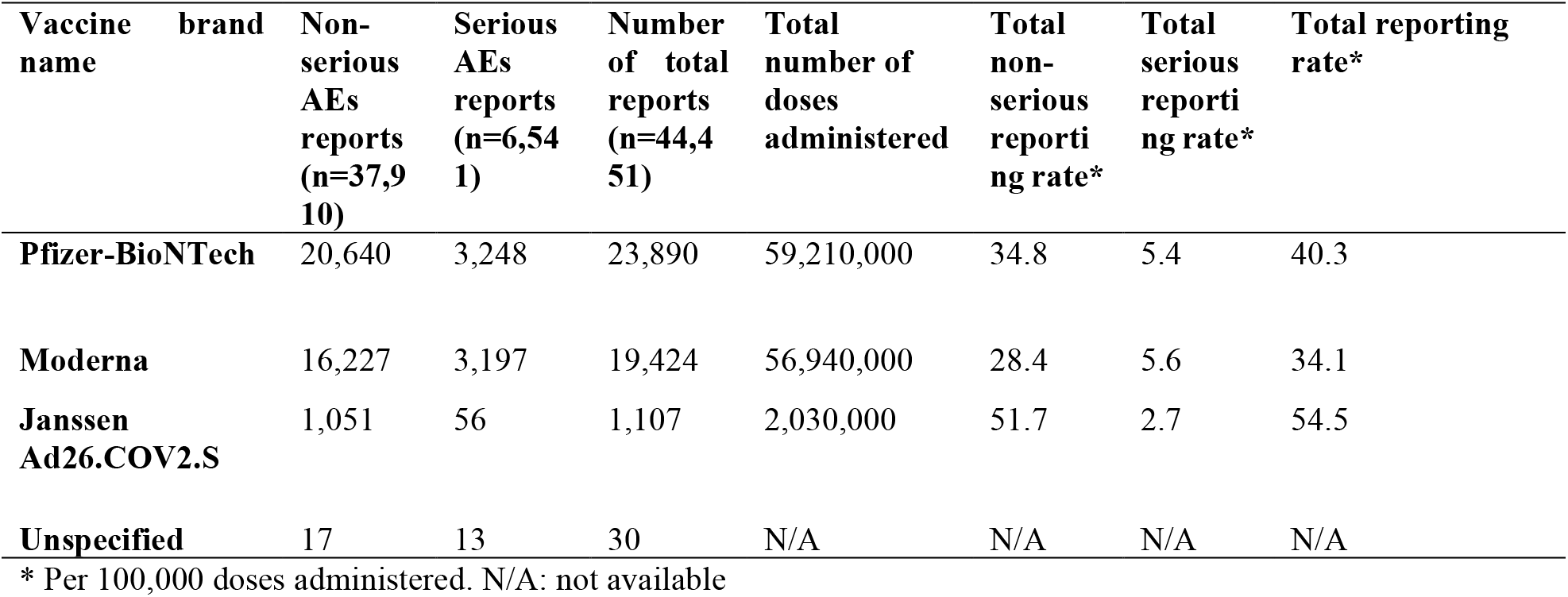
Number and report rate (per 100,000 doses administered) of AES reports per vaccine

Serious AEs were present in 6,514 (14.0%) total AE reports (corresponding to 0.007 % people vaccinated) (Table 3). Most patients with serious AEs were elderly (median: 70 years [range 16-106]). The majority of these serious AEs (4,018 [9.0%]) led to hospitalization (0.004 % of people vaccinated). Life-threatening complications were reported in 431 (0.9%), corresponding to 0.0005% people vaccinated. Anaphylaxis was reported in 326 AE reports (0.73% of AE reports, 0.00038 % people vaccinated), and 130 of 326 (39.8%) of these cases occurred among patients with a documented history of allergy. Thromboembolism was identified in 173 reports (0.3% of AE reports, 0.00002 percent of people vaccinated). Of the 173 cases, 94 (54%) occurred after the vaccination with Moderna, 78 (45%) after Pfizer-BioNTech. Pregnancy-related AEs (spontaneous abortion) were identified in only 64 reports.

**Table 3.**
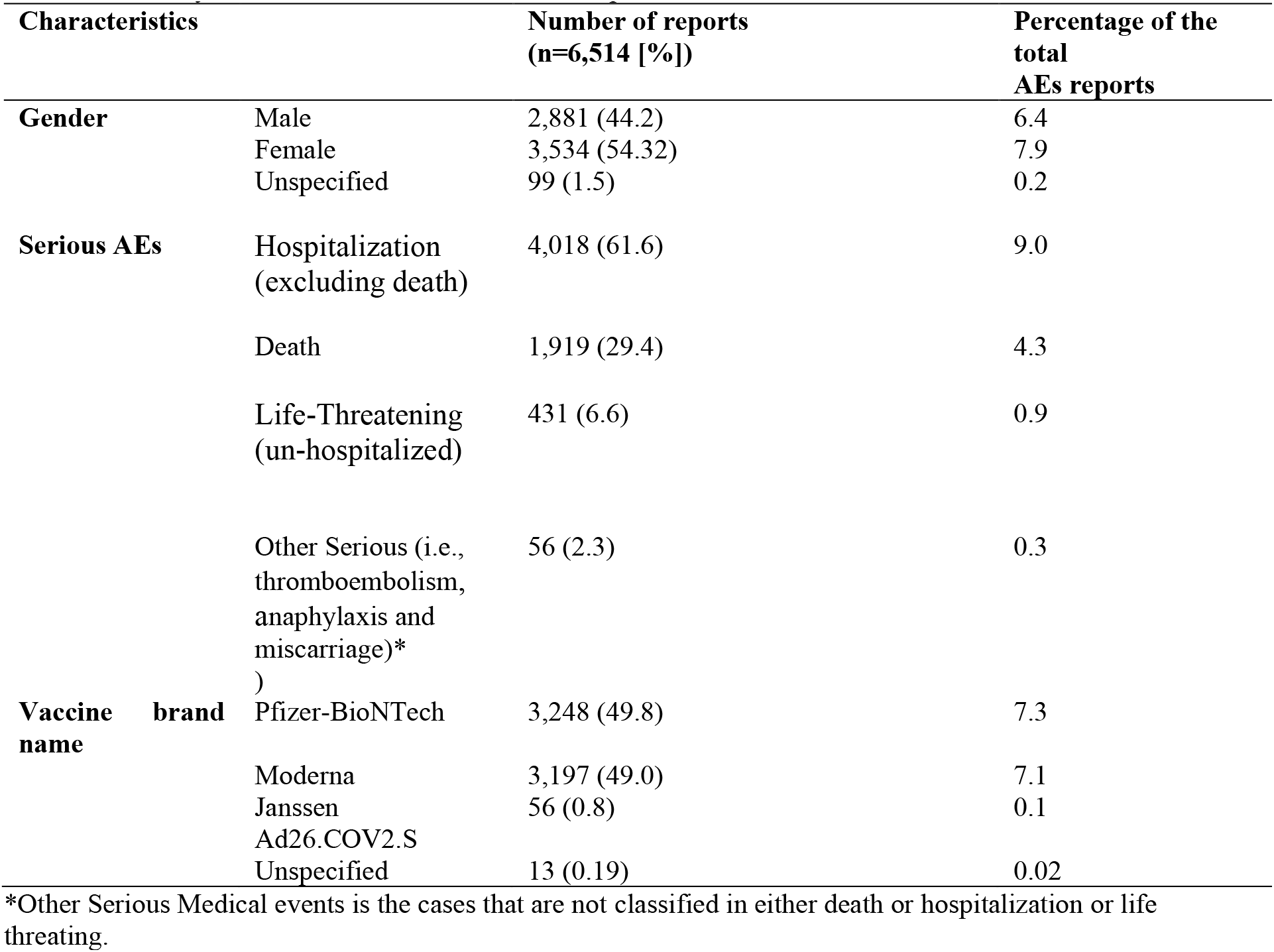
Summary of serious adverse events after receipt of COVID-19 vaccines

Death was identified in 1,919 reports (4.3% of 44,451 AE reports, corresponding to 0.002 % people vaccinated). The median age at death was 80 years (range 18-106 years), and more than one-third (38.4 %) of patients had a history of a CVD. The most commonly reported cause of death was death due to a cardiovascular disease (including cardiac arrest) in 273 of 1,919 (6.0%) of death cases. Fifty-seven patients were already in hospice and 119 had COVID-19 infection. Of the total death reports, 997 (52.0%) were reported after the vaccination with Moderna, 899 (46.8%) after Pfizer-BioNTech, 16 (0.8%) after Janssen Ad26.COV2.S (Table 4). The numbers of death cases between Moderna vs. Pfizer-BioNTech were comparable (1.07; 95% confidence interval 0.86 to 1.33).

**Table 4.**
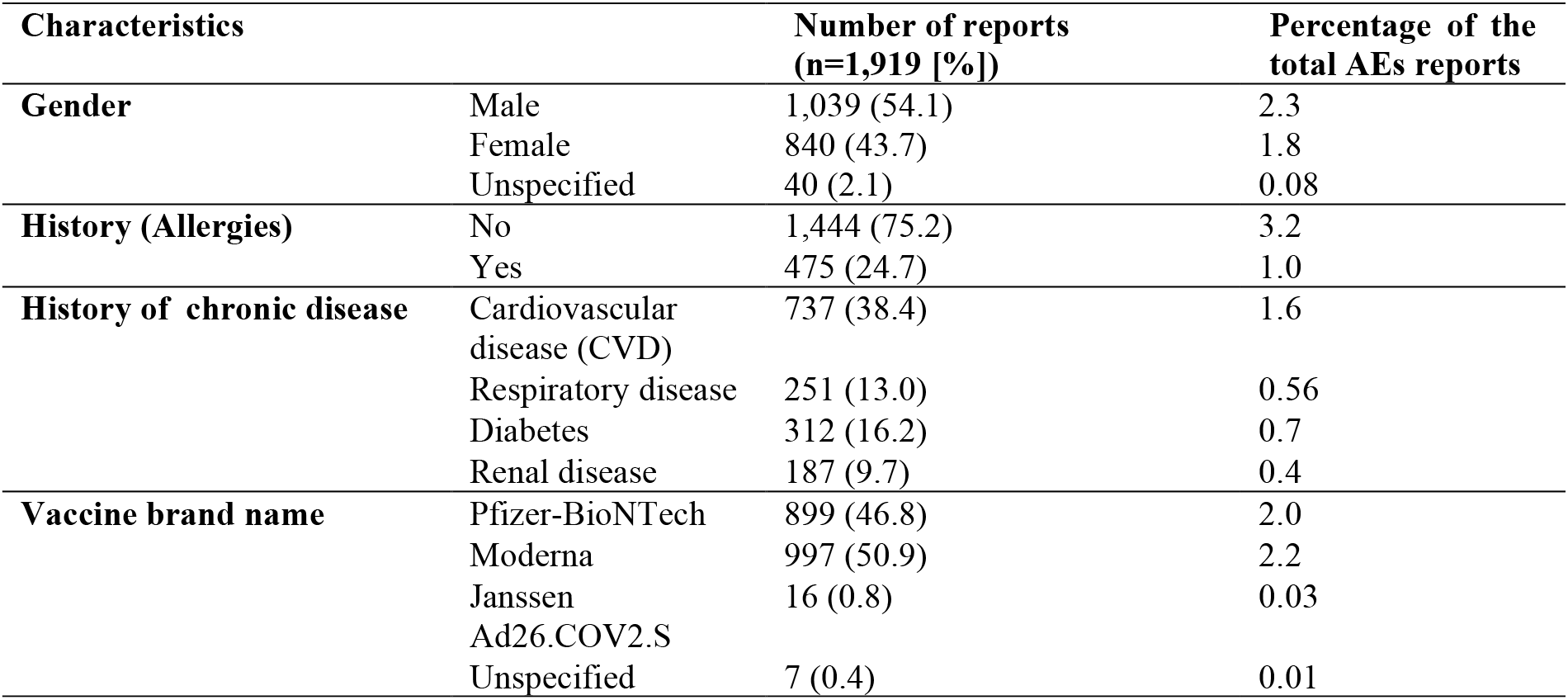
Characteristics of the death cases after receipt of COVID-19 vaccines

## 4. Discussion

Our study showed that most of the reported AEs among those who received the three approved COVID-19 vaccines in the US were non-serious. The study also showed no emerging trends in unknown serious AEs (the most commonly reported serious AEs were reported were hospitalization, followed by death and life threating). Finally, death cases between Moderna vs. Pfizer-BioNTechCOVID-19 vaccines.

The proportion/report rate of AEs observed in our study are consistent with the findings of the previous studies (from a population-level and rate perspectives) that used a spontaneous-reporting database (both in the U.S. and internationally [10]. For example, the percentage of the total reported AEs among the vaccinated population (0.05%) was similar to the observed one in previous VAERS-based PMS assessment of the first month post-authorization (0.05%) [10]. Additionally, the reporting rate in our study (37.5 cases per 100,000 doses administered) was similar to that reported in a Canadian analysis (31.0 reports per 100,000 doses of all administered up to 21 May 2021) [19]. The higher reporting percentage among the females in our study was also observed in previous studies [10,19–21]. This difference has been attributed to several potential risk factors such as (e.g. genetic differences and the higher tendency to report AEs among the female group) [19,22].

The percentage of serious adverse events (14.6%) in our study is higher than that of a previous VAERS-based study (9.2%) [10]. The reporting rate of anaphylactic cases was lower in comparison with a previous study (2.7 vs. 4.5 reported cases per million doses administered) [10]. The number of spontaneous abortion cases we observed in our study (64 reports) is slightly higher than the 46 cases observed in a previous analysis of COVID-19 vaccine PMS data among the pregnant U.S. population, which covered the period up to 28 February 2021 (14). The percentage of the overall of death cases (4.3%) in our study (1,919 of 44,451) was higher than that of a previous VAERS-based study (1.3%) [10]. The median age at death was 80 years (most with co-morbidities), and a similar finding was reported in other countries [23]. We could not find any differences in the number of death cases between the most commonly used COVID-19 vaccines in the U.S. (i.e. Pfizer vs. Moderna).

The number of the retrieved AE reports allowed us to extend the PMS assessment to diverse US populations and to populations that were not covered in the pre-marketing clinical development programs of the three approved vaccines. However, our analyses were conducted in the context of the spontaneous (unsolicited) AE reporting passive framework of VAERS. This precluded us from running all typical risk-, rate-or hazard-based analyses (i.e. having the actual population of the study as a denominator). It also precluded us from running several comparative analyses among the three vaccines adjusting for the relevant measured confounders. Future real-world studies using electronic health records (EHRs) would be more informative in the post-marketing safety assessment of these vaccines.

## 5. Conclusion

Our study showed that the vast majority of the PMS AEs in the U.S. were non-serious and comparable to the findings of the Phase III trials. The study also showed no emerging trends in unknown serious AEs. Finally, we did not find any differences in the number of reported death cases between Moderna vs. Pfizer-BioNTechCOVID-19 vaccines. Future large-scale, real-world studies are needed to confirm the findings of this study.

## Data Availability

Data used in this study are available and accessible through VAERS public domain ().

https://vaers.hhs.gov/

## Declarations

### Availability of data and material

Data used in this study are available and accessible through VAERS public domain (http://vaers.hhs.gov/index).

### Code availability

All data were coded using Microsoft Excel® 2013 (Microsoft Corporation, Redmond, WA, USA) and analyzed using RStudio Version 1.4.1103.

### Author contributions

The study was design and co-supervised by Thamir Alshammari and Turki Althunian. Omar Albalawi and Maha Alomran contributed to the conception and design of the study. Maha Alomran, Omar Albalaw and Ghada Alsagry completed the data extraction, linkage Omar Albalaw and Maha Alomran conducted the descriptive statistical analyses. Turki Althunian conducted the regression analysis of mortality data. Omar Albalawi drafted the manuscript, and Turki Althunian supervised the writing process and made a substantial contribution to editing the manuscript. The final version was reviewed by Thamir Alshammari. All authors were reviewed approved the final version of the manuscript.

### Ethics approval

This study was conducted retrospectively from de-identified data obtained for pharmacovigilance and clinical purposes (publicly available). Thus, no ethical approval was needed.

### Consent to participate

Not applicable.

### Consent for publication

Not applicable.

### Disclaimer

The views expressed in this paper are those of the author(s) and not do not necessarily reflect those of the Saudi Food and Drug Authority or its stakeholders. Guaranteeing the accuracy and the validity of the data is a sole responsibility of the research team.

